# Construction of a whole-brain segmentation pipeline for 0.35T neonatal MRI

**DOI:** 10.64898/2026.09.23.26363741

**Authors:** Zhexian Sun, Jian Huang, Xi-Nian Zuo, Hongjian He, Gang Yu

## Abstract

**Background:** Whole-brain segmentation of neonatal low-field MRI is needed for regional volumetric analysis, but established segmentation systems are designed around different field strengths, contrasts, and age ranges.

**Methods:** We first applied complete end-to-end pipelines, including FreeSurfer, FSL-based processing, Infant FreeSurfer, FastSurfer, BrainSuite, SPM12/CAT12, iBEAT v2, the dHCP structural pipeline, and M-CRIB-S, directly to 0.35T neonatal T2-weighted images. Because those systems did not produce a complete 47-region output, we next tested released end-to-end AI segmentation models, including SynthSeg, FastSurfer, FastSurferVINN, and QuickNAT. These were highly sensitive to input format, contrast, voxel geometry, and the high-field training domain. We then decomposed the task into skull stripping, registration, super-resolution, atlas propagation, and tissue refinement and tested the available components individually.

**Results:** No complete end-to-end pipeline or released AI model delivered a usable 47-region segmentation in direct application. Stage-wise testing showed that preprocessing, reconstruction, registration, atlas propagation, and expectation-maximization each solved only part of the problem. Their final assembly produced complete 47-region outputs across the neonatal cohort.

**Conclusions:** Direct application of established pipelines was insufficient for 0.35T neonatal T2-weighted MRI. A stage-wise combination of compatible components provided the workable solution.

## 1. Introduction

The first months after birth contain a period of exceptionally rapid and regionally heterogeneous brain growth. Volumetric MRI can resolve changes in cortical, subcortical, ventricular, and white-matter structures that are not captured reliably by head circumference or cranial ultrasound, and regional volumes have been associated with later neurodevelopmental outcomes [1-5]. Reliable whole-brain segmentation is therefore a prerequisite for studying early brain maturation and for constructing clinically interpretable growth references.

Most established structural pipelines were developed for adult or high-field neonatal T1-weighted MRI. FreeSurfer [6] and Infant FreeSurfer [7] use cortical and subcortical priors optimized for those contrasts, while the dHCP pipeline [8] assumes a specialized high-field neonatal acquisition. The M-CRIB atlas [9] provides neonatal anatomical labels, but atlas propagation still depends on registration and subject-specific refinement. These systems are individually mature, but they are not interchangeable with thick-slice 0.35T T2-weighted input.

End-to-end AI segmentation has added a second class of solutions. FastSurfer [10], FastSurferVINN [11], QuickNAT [12], and SynthSeg [13] can produce whole-brain labels without manual feature engineering, while nnU-Net [14] provides a self-configuring training framework. These methods are highly sensitive to the input distribution, including contrast, orientation, voxel spacing, intensity normalization, and skull coverage. Applying a model outside its training domain can produce a completed inference while still returning anatomically implausible labels [15].

Low-field MRI offers a practical alternative for neonatal imaging because it is quieter, more portable, and easier to integrate into routine care [16-17]. Its benefits are accompanied by lower signal-to-noise ratio, thick slices, and scanner-specific contrast, all of which weaken the assumptions of high-field segmentation systems. A workable method therefore has to address image formation, spatial alignment, anatomical priors, and tissue classification as connected but inspectable stages.

We first tested whether complete end-to-end pipelines could solve the task directly. We then evaluated released AI segmentation models and their sensitivity to input format and training domain. After both approaches failed to deliver a coherent 47-region output, we decomposed the problem into skull stripping, super-resolution, registration, atlas propagation, and tissue refinement, tested the available compartments within each stage, and assembled the final analysis pipeline from the components that completed their assigned task.

## 2. Data and acquisition

This study used the published 100-infant low-field MRI dataset from the Scientific Data resource, which contains 54 female and 46 male term-born infants aged 1 to 70 days. The release provides raw NIfTI images, manual brain masks generated in MITK, N4-corrected images and bias fields, subject-to-M-CRIB initial transforms, BIDS metadata, and atlas-based segmentation derivatives for ten selected regions [16]. Table 1 summarizes the study cohort.

**Table 1.** Characteristics of the 100-infant public study dataset.

| Characteristic | Public testing resource |
| --- | --- |
| Subjects | 100 infants (54 female, 46 male) |
| Age | 1-70 days chronological age |
| Scanner | 0.35T NIDO (Jiangsu Lici) |
| Acquisition | T2-weighted FSE; 0.82/0.89 mm in-plane; 6-7 mm slice thickness |
| Reference masks | Manual MITK segmentation |
| Segmentation derivatives | ANTs and Draw-EM |
| Regions | Ten selected ROIs |
| Access | Science Data Bank data-use agreement |

The study dataset includes term-born infants without visible radiological abnormality. Manual masks were generated in MITK and atlas-derived outputs were produced with ANTs and Draw-EM. DUA, data-use agreement; LF-MRI, low-field MRI; ROI, region of interest.

### 2.1. Literature search and model selection

We searched Crossref and the original publications for whole-brain segmentation methods, neonatal brain segmentation, learned structural segmentation, and atlas-based neonatal pipelines. Priority was given to methods that were widely used, published in established neuroimaging or methods venues, and supported by a public implementation or pretrained weights. Methods without released weights were retained as trainable frameworks rather than evaluated as ready end-to-end models.

The search covered classical structural pipelines, released deep-learning models, trainable segmentation frameworks, registration methods, brain-extraction tools, and neonatal atlas resources. A method was eligible for direct comparison if it could accept structural MRI input and produce a whole-brain or regional segmentation. Methods producing only a brain mask or registration transform were assigned to the corresponding stage-wise comparison.

## 3. First wave: direct application of complete pipelines

The first wave evaluated complete pipelines rather than isolated algorithms. FreeSurfer included recon-all and its internal extraction and labeling components, including SynthStrip and SynthSeg where available [6,13,18]. FSL was tested as a combined workflow assembled from BET2, FLIRT, FNIRT, and FAST [19]. ANTs was evaluated as a multistage cortical and label-fusion workflow [20]. We also tested Infant FreeSurfer [7], FastSurfer [10], BrainSuite, SPM12/CAT12, iBEAT v2, the dHCP structural pipeline [8], and M-CRIB-S. Table 2 summarizes the tested pipelines and their observed failure modes.

**Table 2.** Direct application of complete whole-brain segmentation pipelines.

| Pipeline | Direct outcome | Main failure mode |
| --- | --- | --- |
| FSL-derived workflow | Extraction, registration, or tissue maps only | No complete 47-region output |
| FreeSurfer | No coherent neonatal LF-T2 segmentation | T1/adult assumptions; internal components do not compensate |
| ANTs-derived workflow | Modular transforms and labels | Needs neonatal atlas priors and refinement |
| Infant FreeSurfer | Incomplete LF-T2 output | High-field T1 domain |
| FastSurfer | Wrong label inventory | High-field T1 domain |
| BrainSuite | No usable 47-region result | Structural high-field assumptions |
| SPM12/CAT12 | Tissue maps rather than target regions | Label hierarchy mismatch |
| iBEAT v2 | No complete target-label output | High-field infant domain |
| dHCP pipeline | Failed under LF input assumptions | Specialized high-field dependencies |
| M-CRIB-S | Atlas alignment without complete LF result | Acquisition and workflow mismatch |

Each complete pipeline was applied directly to 0.35T neonatal T2-weighted MRI using its standard or recommended workflow. Direct outcome describes the best available output before any project-specific adaptation. T1W, T1-weighted.

Nearly all complete pipelines failed in direct application. Failure was expressed as an incomplete output, a label inventory mismatch, an anatomically implausible segmentation, or dependence on high-field and T1-specific priors. This established that the problem required decomposition rather than substitution of one complete pipeline for another.

## 4. Intermediate wave: end-to-end AI whole-brain segmentation models

We next tested released end-to-end AI whole-brain segmentation models that are widely used in high-impact structural neuroimaging. These included SynthSeg, FastSurfer, FastSurferVINN, and QuickNAT [10-13]. We also evaluated trainable frameworks, including nnU-Net and MONAI [14], but these required neonatal LF-specific labels and validated weights before they could be applied as complete models. Table 3 lists the tested AI models and trainable frameworks.

**Table 3.** Released end-to-end AI models and trainable segmentation frameworks.

| Model or framework | Publication | Direct LF-T2 outcome | Failure mechanism |
| --- | --- | --- | --- |
| SynthSeg | Medical Image Analysis, 2023 | Boundary instability and label mismatch | High-field training distribution and voxel-geometry mismatch |
| FastSurfer | NeuroImage, 2020 | No coherent target label inventory | T1, resolution, and age-domain mismatch |
| FastSurferVINN | NeuroImage, 2022 | Failed on 6-7 mm LF-T2 input | High-resolution T1 assumptions |
| QuickNAT | NeuroImage, 2019 | Anatomically implausible labels | Adult high-field T1 training and label-space mismatch |
| nnU-Net | Nature Methods, 2020 | No pretrained LF neonatal model | Requires task-specific labelled training data |
| MONAI | Medical imaging framework | No ready whole-brain LF neonatal model | Framework rather than a released model |

Models with released weights were tested as complete inference pipelines. Trainable frameworks were classified separately because they require task-specific labels and validated weights. LF-T2, low-field T2-weighted.

The AI models were particularly sensitive to input format. Orientation, voxel spacing, intensity normalization, skull coverage, and contrast had to match the training distribution for the models to behave as intended. Several models completed inference but returned anatomically implausible labels, which showed that execution alone is not evidence of successful segmentation.

## 5. Second wave: stage-wise component testing

The second wave broke the segmentation problem into five operational stages: skull stripping, registration, super-resolution, atlas propagation, and tissue/regional refinement. Component methods were compared only within their stage, preventing extraction, registration, and segmentation tools from being treated as competing end-to-end pipelines. Table 4 summarizes the stage-wise candidates, limitations, and selected components.

**Table 4.** Stage-wise component assessment and final component selection.

| Stage | Tested components | Limitation | Selected component |
| --- | --- | --- | --- |
| Skull stripping | BET2, robust BET, SynthStrip, HD-BET, ROBEX, 3dSkullStrip, ANTs BrainExtraction | No extractor generalized across all PMA or motion levels | Best-performing mask stage within the assembled workflow |
| Registration | FLIRT, FNIRT, ANTs SyN/QuickSyN, NiftyReg, Greedy, elastix, 3dAllineate, 3dQwarp | Unstable transforms after thick-slice initialization | ANTs rigid-affine-SyN |
| Super-resolution | SCSRN and interpolation | Interpolation did not recover through-plane structure | SCSRN |
| Atlas propagation | M-CRIB, UNC 4D atlas, dHCP atlas, MALP-EM, Joint Label Fusion | Atlas labels required subject-specific refinement | M-CRIB with inverse propagation |
| Tissue/regional refinement | FAST, mri_ca_label, SynthSeg, SPM12, CAT12, iBEAT, Draw-EM | Tissue classes or unstable regional boundaries | Draw-EM expectation-maximization |
| Learned alternatives | nnU-Net, MONAI, VoxelMorph, DeepReg, TransMorph | No validated neonatal LF-T2 weights | Deferred |

Components were compared only within their assigned pipeline stage. The final assembly combined the selected extractor, registration method, super-resolution model, atlas prior, and tissue-refinement method. PMA, postmenstrual age.

## 6. Final assembled pipeline

The final assembly used N4 bias correction, SCSRN super-resolution, ANTs rigid-affine-SyN registration, inverse M-CRIB atlas and tissue-prior propagation, and Draw-EM expectation-maximization refinement [9,20-22,27]. The assembled workflow generated complete 47-region segmentations across the 100-infant study cohort.

### 6.1. Image preparation and N4 bias correction

The first module normalizes low-frequency intensity variation across the receive field. N4 bias correction estimates a smooth multiplicative field using B-spline regularization, preserving tissue boundaries while reducing intensity drift across the field of view [21]. This stage is necessary before super-resolution and registration because low-field images contain scanner-specific shading that can bias both neural reconstruction and mutual-information registration.

Alternative intensity-preprocessing options, including uncorrected input and simple intensity normalization, were less stable at the image periphery. N4 was retained because it is deterministic, widely used in neonatal structural pipelines, and does not require training data. The output is a corrected T2-weighted image and an estimated bias field, both retained for reproducibility.

### 6.2. Super-resolution with SCSRN

The native 6-7 mm slice thickness introduces large through-plane voxels that are poorly matched to isotropic neonatal atlas labels. SCSRN reconstructs the thick-slice scan onto a 1-mm isotropic grid using structural constraints learned from high-resolution brain images [22]. The module does not create new anatomical regions; its purpose is to provide a stable grid for nonlinear registration and label propagation.

Interpolation-based upsampling, registration of the original thick-slice images, and deep-learning segmentation without reconstruction were tested as alternatives. Interpolation improved voxel spacing without recovering through-plane structure, while direct registration increased boundary misalignment. SCSRN was retained because it improved subsequent registration and provided a consistent geometry for atlas propagation.

### 6.3. Brain-mask selection

Skull stripping reduces the registration search space and prevents non-brain tissue from influencing atlas alignment. BET2, robust BET, SynthStrip [18], HD-BET [23], ROBEX [24], AFNI 3dSkullStrip, and ANTs BrainExtraction were evaluated. Manual MITK brain masks in the public dataset provided an external reference for mask-level comparison.

No extractor generalized across every postmenstrual age, motion severity, and inferior-coverage pattern. The final workflow therefore treats extraction as an inspectable stage rather than a fixed black box. A failed mask can be identified before it propagates into registration and segmentation.

### 6.4. Registration and deformation model

Registration aligns the reconstructed image to the M-CRIB atlas through rigid, affine, and symmetric diffeomorphic transformations. The rigid stage corrects pose, affine registration accounts for global scale and shear, and SyN estimates a topology-preserving nonlinear deformation. Mutual information is used because it does not assume a fixed linear relationship between LF-MRI and atlas-template intensities [20].

FLIRT/FNIRT, NiftyReg [25], Greedy, elastix [26], 3dAllineate, and 3dQwarp were evaluated as alternatives. Several methods produced unstable deformation after thick-slice initialization or required more parameter tuning than could be reproducibly justified. ANTs SyN was retained because it provided a stable multistage transform and preserved anatomical topology.

### 6.5. M-CRIB atlas and prior propagation

The M-CRIB atlas provides a neonatal cortical and subcortical reference with an anatomical inventory closer to the developing brain than adult atlases [9]. The inverse registration transform projects both atlas labels and tissue probability maps into subject space. These priors initialize the final segmentation while retaining age-appropriate anatomical definitions.

The UNC 4D infant atlas, dHCP atlas, Joint Label Fusion, and MALP-EM were also considered. These resources differed in age coverage, acquisition assumptions, or label definitions. M-CRIB was retained because its regional taxonomy and neonatal anatomy matched the intended 47-region output.

### 6.6. Draw-EM expectation-maximization refinement

Atlas priors cannot account for all subject-specific intensity variation. Draw-EM expectation-maximization updates tissue and regional probabilities using the transformed priors and the individual image, producing subject-specific boundaries [27]. This stage converts approximate atlas alignment into a complete regional segmentation.

FAST, mri_ca_label, SynthSeg [13], SPM12, CAT12, and iBEAT outputs were compared. These methods pro uced tissue classes, adult label inventories, or incomplete neonatal regional maps. Draw-EM was retained because it accepts the M-CRIB priors, operates in native subject space, and returns the required regional labels.

### 6.7. Regional output, nomenclature, and quality control

The final module converts each label map into regional volumes and publication-standard anatomical names. Duplicate atlas identifiers are consolidated, and labels are organized into cortical, subcortical, ventricular, cingulate, insular, and white-matter categories. Radiologist review identifies failed masks, unstable registrations, and anatomically implausible boundaries before volumetric analysis.

**Fig. 1.**
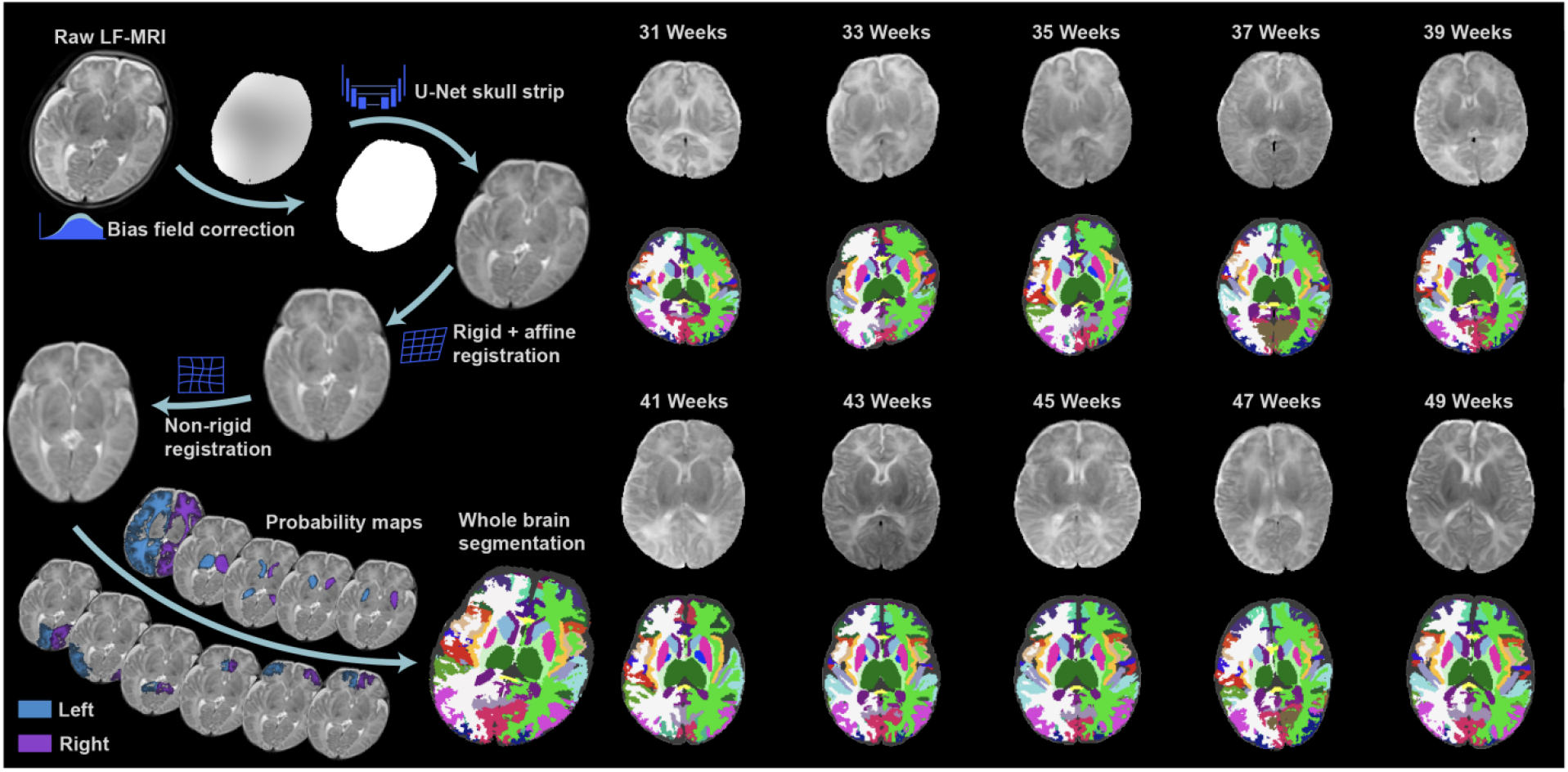
Final assembled segmentation pipeline. Raw low-field MRI is bias corrected, reconstructed to an isotropic grid, registered to the M-CRIB atlas, and refined with atlas-derived tissue priors. Representative whole-brain segmentations from 10 infants spanning 31-49 weeks postmenstrual age are shown.

## 7. Public study dataset

The 100-infant study dataset provides manual brain masks, N4 derivatives, registration inputs, and ten-region atlas- based outputs. It supports evaluation of brain-mask generation, registration consistency, and selected regional volumes. Its ten-region scope defines the boundary of direct regional assessment.

## 8. Discussion

The three-stage design explains why direct application of complete pipelines and released AI models failed. End-to-end systems are optimized for specific contrasts, field strengths, anatomical priors, and output taxonomies. Low-field neonatal T2-weighted MRI violates several assumptions simultaneously.

The AI models were especially sensitive to input-format and domain mismatch. Orientation, voxel spacing, intensity normalization, contrast, and skull coverage had to match the training distribution. Several models completed inference but returned anatomically implausible labels, showing that a successful run does not establish a valid segmentation.

The stage-wise approach also gives the final pipeline a clear performance structure. Each component can be replaced or retested without changing the whole workflow, and a failure in registration can be distinguished from a failure in issue classification. The final assembly is therefore a practical solution to the observed LF-MRI segmentation problem, not merely a concatenation of software packages.

Limitations include the absence of an independent full 47-region reference in the public resource and the need for further quantitative validation of each selected component. The public dataset nevertheless provides an external check for mask generation, registration, and ten-region output.

## 9. Conclusion

Complete established pipelines did not solve segmentation of 0.35T neonatal brain MRI in direct application. Decomposing the task and selecting compatible components produced a workable stage-wise pipeline that generated complete 47-region outputs.

## Data Availability

The external dataset is available through Science Data Bank (DOI https://doi.org/10.57760/sciencedb.o00133.00006) after registration and completion of its data-use agreement.

https://www.scidb.cn/en/detail?dataSetId=325216c8c30d42ee9cc5037e42b74304

## Data and code availability

The external dataset is available through Science Data Bank (DOI 10.57760/sciencedb.o00133.00006) after registration and completion of its data-use agreement. Code and parameter files are available from the corresponding author subject to institutional data-use requirements.

## Ethics approval and consent

The original study was approved by the Medical Ethics Committee of Children’s Hospital, Zhejiang University School of Medicine (IRB/EC 2023-IRB-0287-P-01). Written informed consent was waived because the data were de-identified. Use of the public dataset follows its data-use agreement.

## Funding

This work was partially supported by the National Key R&D Program of China (2023YFC2706400), the National Natural Science Foundation of China (82372036, 62576314), the Zhejiang Provincial Natural Science Foundation of China (LMS26F020001), and the Medical Health Science and Technology Project of Zhejiang Province (2022KY192).

## Competing interests

The authors declare no competing interests.

## Notes

### Author Declarations

Ethics committee/IRB of Children's Hospital, Zhejiang University School of Medicine gave ethical approval for this work.

